# Eicosanoid Inflammatory Mediators Are Robustly Associated with Blood Pressure in the General Population

**DOI:** 10.1101/2020.02.08.20021022

**Authors:** Joonatan Palmu, Jeramie D. Watrous, Kysha Mercader, Aki S. Havulinna, Kim A. Lagerborg, Aaro Salosensaari, Mike Inouye, Martin G. Larson, Jian Rong, Ramachandran S. Vasan, Leo Lahti, Allen Andres, Susan Cheng, Pekka Jousilahti, Veikko Salomaa, Mohit Jain, Teemu J. Niiranen

## Abstract

Epidemiological and animal studies have associated systemic inflammation with blood pressure (BP). However, the mechanistic factors linking inflammation and BP remain unknown. Fatty acid derived eicosanoids serve as mediators of inflammation and have been suggested to also regulate renal vascular tone, peripheral resistance, renin-angiotensin system, and endothelial function. We therefore hypothesize that specific pro- and anti-inflammatory eicosanoids are linked with BP. We studied a population sample of 8099 FINRISK 2002 participants randomly drawn from the Finnish population register (53% women, mean age 48±13 years) and, for external validation, a sample of 2859 Framingham Heart Study (FHS) Offspring study participants (55% women, mean age 66±9 years). Using non-targeted liquid chromatography-mass spectrometry, we profiled 545 distinct high-quality eicosanoids and related oxylipin mediators in plasma. Adjusting for conventional hypertension risk factors, we observed 187 (34%) metabolites that were significantly associated with systolic BP (*P* < Bonferroni-corrected threshold of 0.05/545). We used forward selection linear regression modeling in FINRISK to define a general formula for individual eicosanoid risk score. Individuals of the top risk score quartile in FINRISK had a 9.0 mmHg (95% CI 8.0-10.1) higher systolic BP compared with individuals in the lowest quartile in fully adjusted models. Observed metabolite associations were consistent across FINRISK and FHS. In conclusion, plasma eicosanoids demonstrate strong associations with BP in the general population. As eicosanoid compounds affect numerous physiological processes that are central to BP regulation, they may offer new insights regarding pathogenesis of hypertension, as well as serve as potential targets for therapeutic intervention.

## Introduction

A vast majority of hypertensive patients (>95%) are classified as having primary (essential) hypertension, a heterogeneous condition of hypertension that has no identifiable cause (by definition). Essential hypertension is most likely the consequence of an interaction between genetic factors and environmental factors (e.g., obesity, insulin resistance, sedentary lifestyle, stress, and sodium intake).^1^ Intriguingly, all of the aforementioned factors are also related to chronic low-grade inflammation, underscoring the need to further investigate inflammation as a potential mainstay pathologic mechanism underlying hypertension.^2^

The upstream initiation of inflammatory activity in humans is governed mainly by substrates and products of polyunsaturated fatty acids.^3^ Termed eicosanoids, the small molecule derivatives of arachidonic acid and other polyunsaturated fatty acids serve as both activators and suppressors of systemic inflammatory activity.^3^ Data derived mainly from animal studies suggest that eicosanoid compounds affect renal vascular tone, urine sodium excretion, peripheral resistance, kidney disease, renin-angiotensin-aldosterone system, and endothelial function – factors that are central to blood pressure (BP) regulation itself.^4–9^ Published data have also demonstrated that a few, select eicosanoids, such as 20-hydroxyeicosatetraenoic acid (20-HETE), and genetic polymorphisms that regulate the levels of these eicosanoids, are altered in small samples of individuals and animals with hypertension.^10–12^

Until recently, sensitive methods for detecting and quantifying eicosanoids in large sample sizes were lacking. However, mass spectrometry (MS) based analytics now allow for the rapid large-scale quantification of several hundred upstream eicosanoids in human plasma.^13,14^ Our goal was to gain a more detailed understanding of how upstream inflammatory mediators are related to an individual’s prevalence for hypertension. We quantified a comprehensive panel of >500 distinct high-quality upstream eicosanoids and related oxylipin mediators in FINRISK 2002 (N=8099) and Framingham Heart Study Offspring (FHS Offspring; N=2859) cohort participants using liquid chromatography-MS (LC-MS) and related these eicosanoids and eicosanoid profiles to BP traits.

## Methods

### Cohorts

The FINRISK 2002 study used a random population sample of individuals aged 25– 74 years from six geographical areas of Finland. The sampling was stratified by sex, region and 10-year age group for a population sample of 13500 individuals; the overall participation rate was 65.2% (N=8798). The sampling has been previously described in detail.^15^ Plasma LC-MS was performed successfully on N=8292 participants. After excluding 193 participants with missing covariate data, N=8099 individuals were included in the analyses as the discovery cohort for the present investigation.

The first-generation (i.e. the ‘Original’) cohort of the FHS included a random sample of two thirds of the adult population of Framingham, MA who were enrolled in a longitudinal community-based cohort study in 1948. The FHS Offspring includes 5124 participants – children of the first-generation cohort and their spouses, who have been re-examined every four-to-eight years since the first examination in 1971. The characteristics and study protocol of FHS Offspring cohort have been published.^16^ For this study, we considered N=3002 individuals who participated in the eight examination cycle of FHS Offspring in 2005–2008 and had assays for eicosanoids with LC-MS. After excluding 143 participants with missing covariates, we included N=2859 participants as the replication cohort.

### Ethical Approval

The FINRISK 2002 study was approved by Coordinating Ethics Committee of the Helsinki University Hospital District. FHS Offspring was approved by Boston University Medical Center’s Institutional Review Board. All participants in both studies provided written informed consent. Participants’ consent to publication of information was not required because the participants remain unidentifiable.

### Clinical Evaluation and Definitions

Participants of both cohorts provided a medical history, including information on medication use, and underwent a physical examination and laboratory assessment of cardiovascular risk factors at baseline. The methodologies of these examinations have been described previously in detail.^15,16^ At all examinations, a healthcare professional performed two (FHS Offspring) or three (FINRISK) sequential BP measurements using a mercury column sphygmomanometer on seated participants according to a standardized protocol. We defined the BP at a given examination as the mean of all sequentially measured BP values. We defined hypertension as BP ≥140/90 mmHg or use of antihypertensive medication. Antihypertensive medication use was based on self-report in both studies. We defined pulse pressure as systolic minus diastolic BP and mean arterial pressure as [(2 × diastolic BP) + systolic BP] / 3. We defined body mass index as weight (kg) divided by height (m) squared and current smoking as self-reported daily used of tobacco products. In FINRISK 2002, prevalent diabetes was defined as self-reported diabetes, a previous diagnostic code indicating diabetes in the nationwide Care Register for Health Care (International Statistical Classification of Diseases and Related Health Problems^17^ (ICD) version 10 codes E10-E14, ICD-9 code 250, or ICD-8 code 250), a previous diabetes medication purchase (ATC code A10*) in the nationwide Prescribed Drug Purchase register, or a diabetes medication code in the nationwide Reimbursed Medication Register. In FHS Offspring, prevalent diabetes was defined as a fasting plasma glucose ≥7.0 mmol/l or self-reported use of glucose-lowering medications.

### Eicosanoid profiling

Using a directed non-targeted LC-MS approach in conjunction with computational chemical networking of spectral fragmentation patterns, we identified 545 eicosanoids and related oxylipins in the FINRISK. The methods of plasma eicosanoid profiling using LC-MS have been previously described in detail.^13,14^ Metabolite data were adjusted for technical variation in off-plate pooled plasma samples and in spike-in internal standards. Missing values were replaced with minimum value for each eicosanoid abundance. The six eicosanoids and related oxylipin mediators included in the risk score were matched between FINRISK and FHS by comparing their LC-MS profiles. These metabolites were also identified, if possible, through comparisons with reference standards and online databases.

### Statistical analyses

We normalized eicosanoid abundances using median absolute deviation; we calculated the median of the absolute difference from the median, and used this value to scale all analyte values for a given assay plate. We used linear and logistic regression models to examine the associations between each eicosanoid molecule and BP traits (systolic BP, diastolic BP, pulse pressure, mean arterial pressure, and hypertension). We adjusted for multiple comparisons using Bonferroni correction to minimize the probability of type I error.^18^ Relations between eicosanoids significantly associated with systolic blood arterial pressure were assessed using Spearman correlation and ordered using hierarchical cluster analysis with complete linkage method. We assessed the multivariable association between eicosanoids significantly related to systolic BP using stepwise linear regression modeling with forward selection and a Bonferroni-corrected inclusion threshold of p=0.05/545. For the six eicosanoids that remained in the models, we calculated eicosanoid risk scores according to the formula β_1_X_1_ + β_2_X_2_ + … + β_n_X_n_, with X_n_ denoting the standardized value for the nth eicosanoid abundance, and β_n_ denoting the regression coefficient from the regression model for systolic BP containing the statistically significant eicosanoids.^19^ We assessed the odds of hypertension and increase in systolic BP by 1-SD increases and by quartiles of the risk scores using unadjusted and multivariable adjusted logistic and linear regression models. We replicated these analyses in FHS Offspring using the eicosanoid abundances in FHS and the regression coefficients from FINRISK. We adjusted all analyses for age, sex, BMI, current smoking, diabetes, antihypertensive medication, and mass spectrometry batch. We used R version 3.6.1^20^ for all analyses. The source code for the analyses is openly available at doi:10.5281/zenodo.3604123.^21^

## Results

The characteristics of FINRISK (N=8099, mean age 48.0±13.1 years, 53.1% female) and FHS (N=2859, mean age 66.3±8.9 years, 54.7% women) cohorts are shown in **Table 1**.

**Table 1.** Characteristics of the discovery (FINRISK) and replication (FHS) samples.

| Characteristics | FINRISK 2002 | FHS |
| --- | --- | --- |
| N | 8099 | 2859 |
| Age, y (SD) | 48.0 (13.1) | 66.3 (8.9) |
| Women, N (%) | 4300 (53.1) | 1564 (54.7) |
| BMI, kg/m<sup>2</sup> (SD) | 26.9 (4.7) | 28.3 (5.4) |
| Systolic blood pressure, mmHg (SD) | 135.1 (20.0) | 128.5 (17.2) |
| Diastolic blood pressure, mmHg (SD) | 79.0 (11.3) | 73.4 (10.1) |
| Pulse pressure, mmHg (SD) | 56.1 (16.1) | 55.1 (16.0) |
| Mean arterial pressure, mmHg (SD) | 97.7 (12.7) | 91.8 (10.5) |
| Hypertension, N (%) | 3567 (44.0) | 1673 (58.5) |
| Antihypertensive medication, N (%) | 1177 (14.5) | 1389 (48.6) |
| Current smoker, N (%) | 2097 (25.9) | 256 (9.0) |
| Diabetes mellitus, N (%) | 446 (5.5) | 393 (13.7) |

Continuous variables are presented as mean (standard deviation). Categorical variables reported as absolute and relative frequencies. BMI, body mass index; FHS, Framingham Heart Study.

### Association between Eicosanoids and BP Traits

Of the eicosanoids and related oxylipin mediators, 187 (34.3%) were significantly associated with systolic BP, 124 (22.8%) with diastolic BP, 177 (32.5%) with mean arterial pressure, 161 (29.5%) with pulse pressure, and 155 (28.4%) with hypertension in FINRISK (**Figure 1, Table S1**). We selected systolic BP as our main outcome variable due to its strong association with cardiovascular diseases. We observed 175 (93.6%) positive and 12 (6.4%) negative associations for systolic BP (**Figure 1, Table S1**). The heatmaps of pairwise correlations for the 187 metabolites related to systolic BP are shown in **Figure 2** and **Figure S1**. This analysis revealed strong overall correlations, but only minor clustering of the eicosanoids.

**Figure 1.**
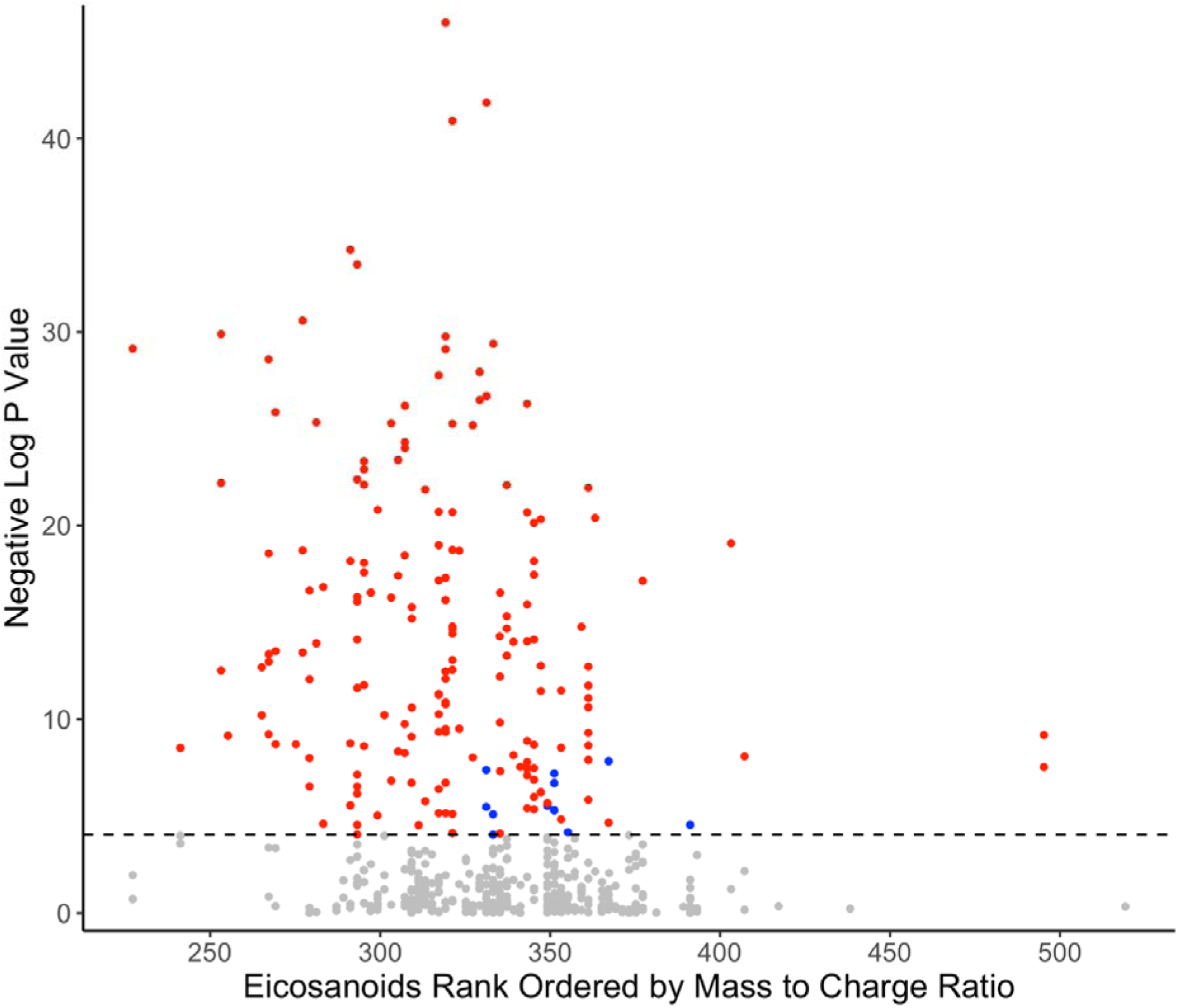
Manhattan plot for associations between metabolites and systolic blood pressure in FINRISK 2002. A significant association was observed for 187 of the 545 eicosanoids. Positive correlations are denoted in red, negative in blue and insignificant in gray. Eicosanoids are ordered by the value of mass to charge ratio (m/z). Dashed line represents the Bonferroni corrected (p=0.05/545) level of significance. Analyses are adjusted for age, sex, BMI, current smoking, diabetes, antihypertensive medication, and batch.

**Figure 2.**
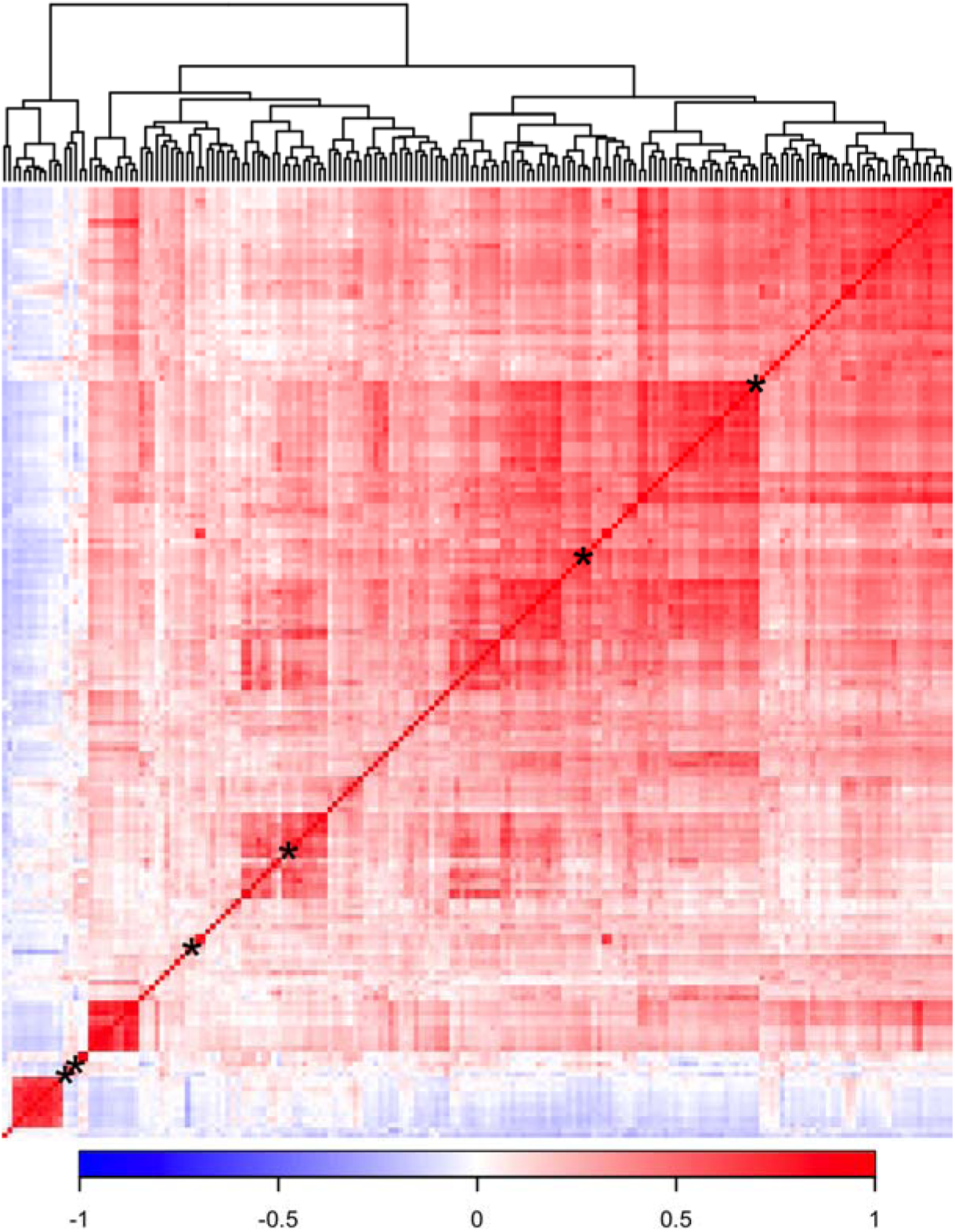
Correlation matrix for the 187 plasma metabolites related to systolic blood pressure in FINRISK 2002. Relations between eicosanoids were calculated using Spearman correlation and ordered using hierarchical cluster analysis with complete linkage method. Only eicosanoids related to systolic blood pressure were included in the correlation matrix. Asterisk denotes position of the six metabolites in our eicosanoid risk score (**Figure 3**).

### Independent Determinants of BP and Hypertension

We used forward selection linear regression modeling with a Bonferroni-corrected inclusion threshold to define a set of metabolites that was independently associated with systolic BP. In FINRISK, these six metabolites were 11-dehydro-2,3-dinor thromboxane B_2_ (TXB_2_), 12-hydroxyheptadecatrienoic acid (12-HHTrE), 265.1809/3.57 (putative eicosanoid), 295.2279/4.89 (putative eicosanoid), 319.2280/5.67 (unknown), and adrenic acid (**Table S2)**. Two of these six metabolites could not be detected in FHS plasma samples (11-dehydro-2,3-dinor-TXB_2_ and 295.2279/4.89). Comparing single metabolite associations, adjusted for relevant covariates, demonstrated that effect sizes between the metabolites were highly consistent across the two cohorts (**Figure 3, Table S3**).

**Figure 3.**
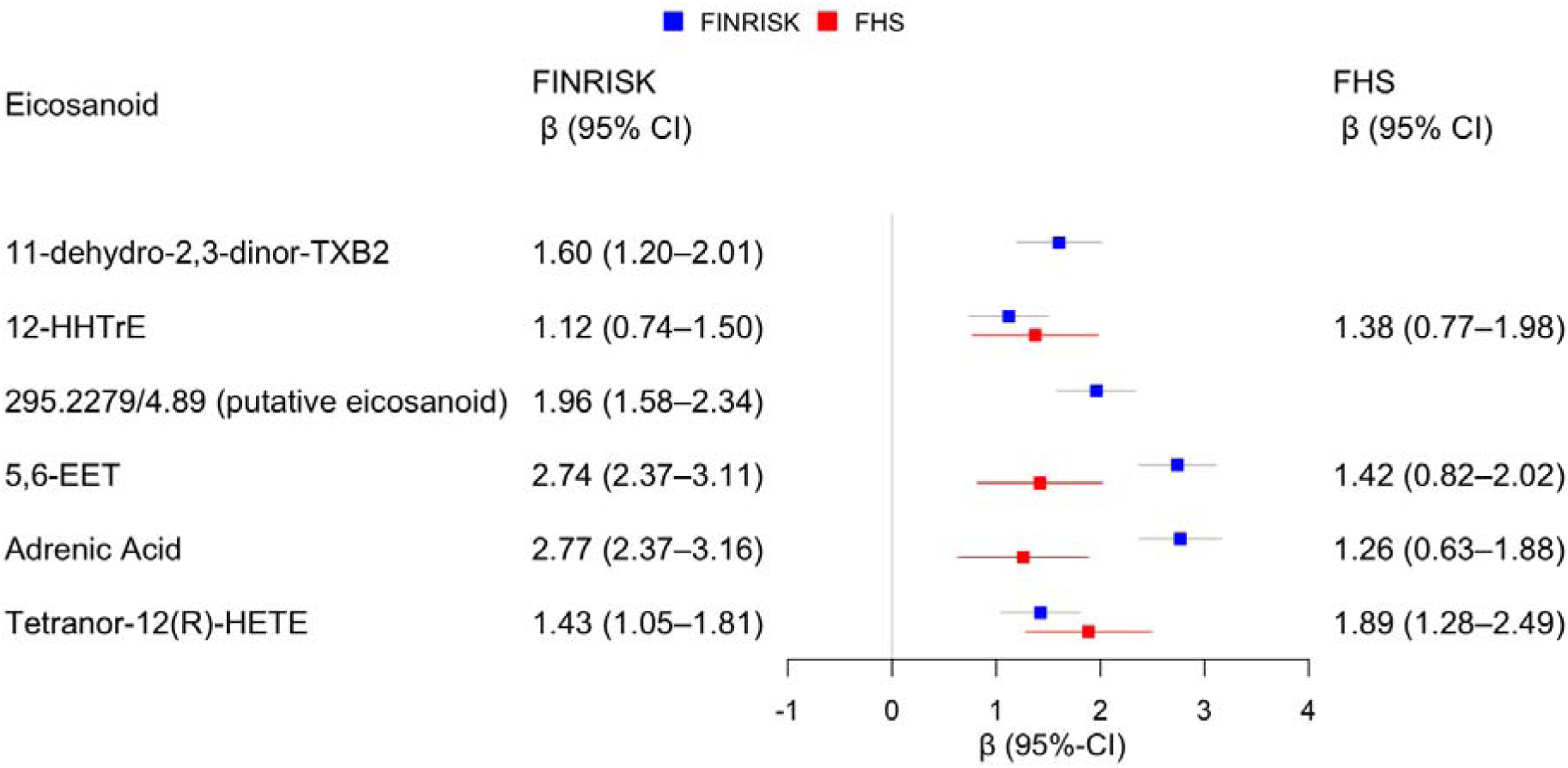
The associations between a subset of six metabolites and systolic BP in FINRISK and replication of results in FHS. The beta coefficients are for the association between 1-SD increase in metabolite concentration and the absolute change of systolic blood pressure (mmHg) in the two study cohorts. All models were adjusted for age, sex, BMI, current smoking, diabetes, antihypertensive medication, and batch. Two of the six eicosanoids observed in FINRISK were not observed in FHS plasma samples. FHS, Framingham Heart Study; HHTrE, hydroxyheptadecatrenoic acid; TXB2, thromboxane B2; HETE, hexadecatrienoic acid.

### Eicosanoid risk score

We defined an eicosanoid risk score using the effect sizes in FINRISK for the six previously mentioned metabolites (**Table S2**). The abundances of the two non-detected metabolites in FHS were treated as zero values. Individuals in the top risk quartile had 9.0 mmHg (95% CI 8.0-10.1 mmHg) higher systolic BP in FINRISK and 6.8 mmHg (95% CI 5.1-8.5 mmHg) in FHS compared with individuals in the lowest quartile (**Figure 4, Figure 5, Table S4, Table S5)**. The odds for hypertension were respectively 2.3 (95% CI 2.0–2.6) and 2.0 (95% CI 1.6-2.5) for the top quartile compared to the lowest quartile (**Figure 4, Figure 5)**. These associations were consistent across FINRISK and FHS.

**Figure 4.**
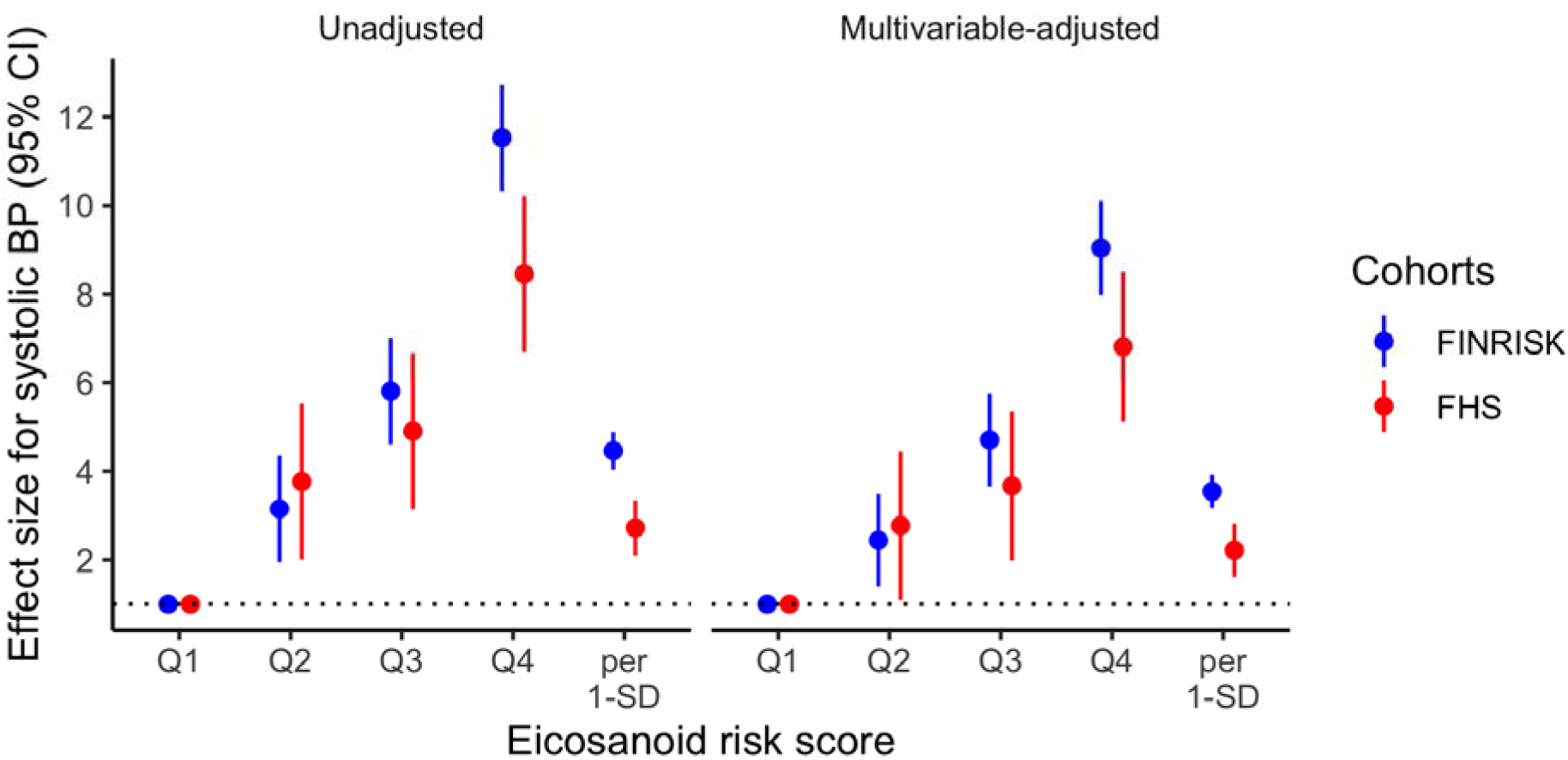
The association between the risk score and systolic blood pressure in FINRISK and FHS. We calculated eicosanoid risk score for each participant according to the formula β_1_X_1_ + β_2_X_2_ + … + β_n_X_n_, with X_n_ denoting the standardized value for the nth eicosanoid abundance, and β_n_ denoting the regression coefficient from the regression model containing the indicated eicosanoids. Analyses are adjusted for age, sex, BMI, current smoking, diabetes and antihypertensive medication and batch. FHS, Framingham Heart Study; BP, blood pressure.

**Figure 5.**
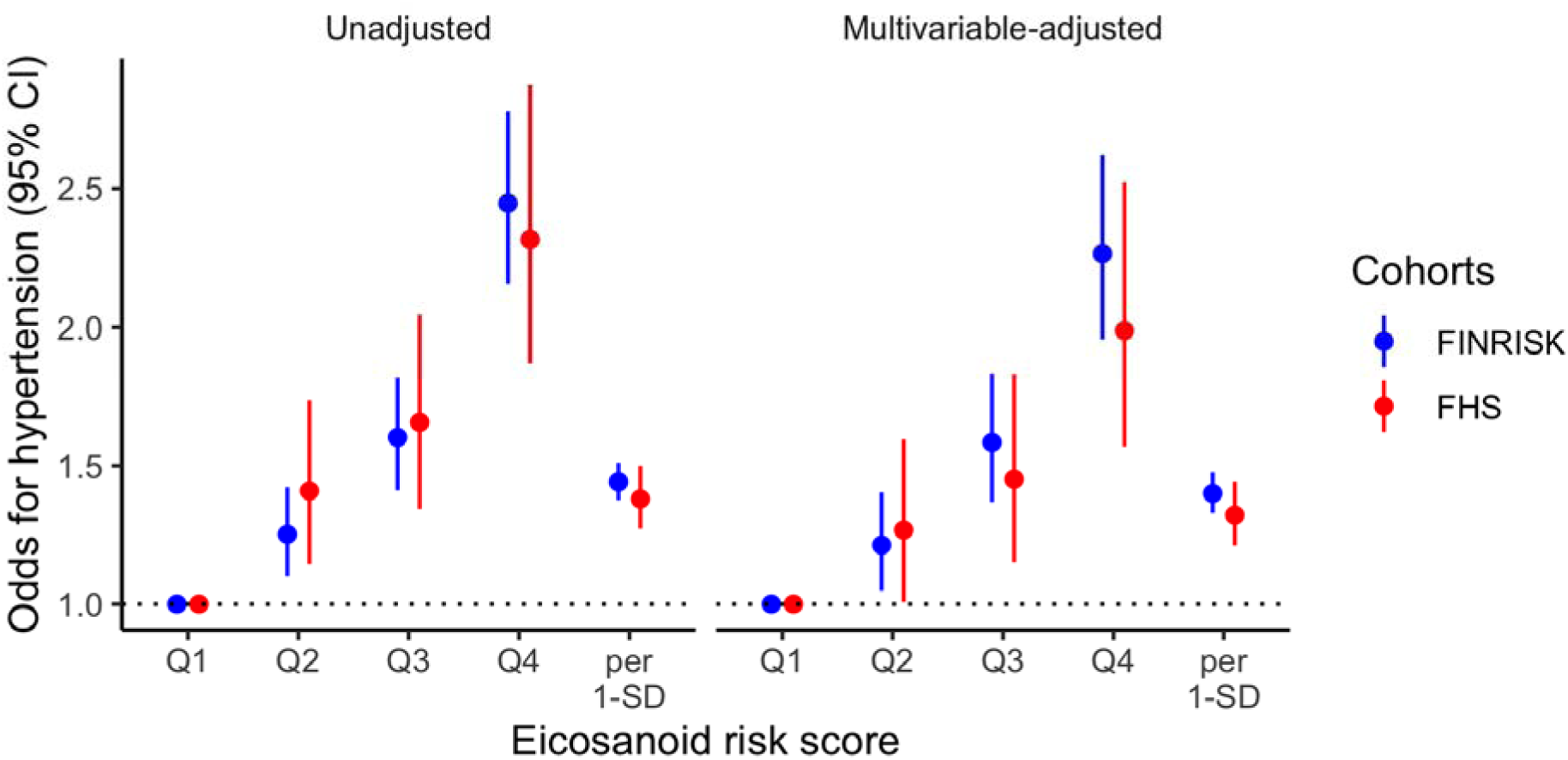
The association between the risk score and hypertension in FINRISK and FHS. We calculated eicosanoid risk score for each participant according to the formula β_1_X_1_ + β_2_X_2_ + … + β_n_X_n_ with X_n_ denoting the standardized value for the nth eicosanoid abundance, and β_n_ denoting the regression coefficient from the regression model in FINRISK containing the indicated eicosanoids. Multivariable analyses are adjusted for age, sex, BMI, current smoking, diabetes and antihypertensive medication and batch. FHS, Framingham Heart Study; BP, blood pressure.

## Discussion

Using a directed non-targeted LC-MS approach in well-phenotyped, large community-based cohorts, we identified 187 eicosanoids and related oxylipins that were associated with systolic BP. FINRISK 2002 participants in the top quartile of the eicosanoid risk score had 9.0 mmHg greater systolic BP and a >2-fold odds of hypertension, compared with individuals in the lowest quartile. These findings were replicated in the FHS Offspring participants.

The upstream initiation of inflammatory activity in humans is governed mainly by substrates and products of polyunsaturated ω-3 and ω-6 20-carbon fatty acids.^7,22^ The small molecule derivatives of arachidonic acid and other polyunsaturated fatty acids, termed eicosanoids, serve as both activators and suppressors of systemic inflammatory activity.^23,24^ The majority of research on systemic inflammation in humans has focused on downstream markers of inflammatory activity such as cytokines and acute phase reactants. Recent work by us and others has shown that chronic elevation in these downstream markers is associated with a variety of cardiovascular disease risk traits and outcomes.^25,26^ In particular, several cross-sectional and prospective studies in humans have found an association of plasma concentrations of downstream low-grade inflammation markers, such as interleukin-6, intercellular adhesion molecule-1, C-reactive protein, and tumor necrosis factor alpha, with arterial stiffness and hypertension.^27–33^. Although downstream markers of inflammation are associated with hypertension and a variety of cardiovascular disease outcomes, evidence for a clinically important, causal role of these biomarkers has been mixed.^34^ In addition, despite inflammation being pivotal in the development of atherosclerosis and certain medications with anti-inflammatory properties clearly reduce cardiovascular disease risk, the extent to which any given inflammatory pathway warrants attention as a direct putative target for therapy is unknown.^35^ Such results have now led experts to suggest that, where inflammation is concerned, causal factors may be upstream.^34^

This study is the first to comprehensively examine the association between eicosanoids and BP in humans. Prior studies with study samples consisting of tens of hypertensive subjects with a panel of a few, mainly cytochrome P450 pathway eicosanoids have demonstrated that eicosanoids, in general, affect regulation of renal function, vascular tone, and the development of hypertension.^12,36–39^ Our results from a large, population-based sample demonstrate that a large number of eicosanoid species are related to BP in both a positive and negative way. In addition, we demonstrate that a distinct eicosanoid score is related to a >2-fold odds of hypertension.

Eicosanoids are metabolized via three general pathways that involve that involve cytochrome P450 monooxygenases, cyclooxygenases, and lipooxygenases. Several of the identified metabolites that remained in the six-eicosanoid risk score are members of these pathways. The cytochrome P450 pathway metabolizes arachidonic acid to several eicosanoids, including 20-HETE and EETs.^8^ These metabolites are critical in BP regulation and also provide cardio- and renoprotective effects in chronic kidney disease.^8^ Cyclooxygenase-derived prostanoids, which include 11-dehydro-2,3-dinor-TXB_2_ identified in our study, modulate renal vasodilatation, natriuretic mediator excretion, and BP during high salt diet.^40–42^ 11-dehydro-2,3-dinor-TXB_2_, has also been associated in clinical studies with thrombogenesis, pregnancy-induced hypertension, and cardiovascular events.^43–45^ Another metabolite that was included in our six-eicosanoid risk score, 12-HHTrE, a product of arachidonic acid metabolism through the 5-lipoxygenase pathway and nonenzymatic degradation product of prostanoids thromboxane A2 and prostaglandin H2, has vasoconstrictor effects.^46,47^ In addition to eicosanoid pathway products, adrenic acid was also included in our risk score. Adrenic acid is a polyunsaturated 22-carbon fatty acid and mainly a substrate for eicosanoid production.^48^ Given the findings from our study and from previous experimental trials, these results provide a strong biological basis for how eicosanoids could affect human BP regulation through several different mechanisms.

Our study has several strengths, such as unselected population sample, external replication of our results, and assays of a large number of eicosanoids. However, our results must be interpreted in the context of potential limitations. First, LC-MS is a highly sensitive method for assessing circulating metabolites. Particularly, of the six metabolites that remained in the final forward-selection regression model in FINRISK, only four were observed in FHS samples. This could in part be explained by the between-cohort age and smoking disparities. Second, the unambiguous metabolite classification and identification is still a challenge in high-throughput LC-MS. However, we have previously demonstrated that these signals are highly consistent with known and putative eicosanoids and related oxylipins in human plasma.^14^ Finally, due to the cross-sectional and observational nature of our study, no causal inferences can be made.

### Perspectives

Plasma eicosanoids demonstrate strong associations with BP in the general population and differences between eicosanoid profiles are observed between normotensive versus hypertensive participants. Intriguingly, although most of the associations were positive (harmful species), we observed protective molecules as well. Thus, eicosanoids may offer new insights regarding pathogenesis of hypertension and related adverse outcomes, as well as serve as potential targets for therapeutic intervention.

## Data Availability

All data is available at THL Biobank (https://thl.fi/en/web/thl-biobank) except for LQ/MS data that is being prepared for publication.

## Acknowledgments

We thank the participants and staff of the FINRISK 2002 and FHS.

## Novelty and Significance

- **What is New**
  - Fatty acid-derived eicosanoids serve as mediators of inflammation and have been suggested to regulate renal vascular tone, peripheral resistance, renin-angiotensin system, and endothelial function.
  - We assayed a comprehensive panel of >500 distinct high-quality eicosanoids and related oxylipin mediators in community-based samples of >10 000 individuals using liquid chromatography-mass spectrometry and relate these eicosanoids and eicosanoid profiles to blood pressure traits.
- **What is relevant**
  - We observed that 187 (34%) eicosanoids and related oxylipin mediators were significantly associated with systolic blood pressure.
  - Individuals in the top quartile of a six-metabolite risk score had a 9.0 mmHg higher systolic blood pressure and 2-fold greater odds of hypertension compared to individuals in the bottom quartile.

## Summary

As eicosanoid species affect numerous physiological processes that are central to BP regulation, they may offer new insights regarding pathogenesis of hypertension, as well as serve as potential new targets for therapeutic intervention.

